# Transfer learning to detect COVID-19 automatically from X-ray images, using convolutional neural networks

**DOI:** 10.1101/2020.08.25.20182170

**Authors:** Mundher Taresh, Ningbo Zhu, Talal Ahmed Ali Ali

## Abstract

Novel coronavirus pneumonia (COVID-19) is a contagious disease that has already caused thousands of deaths and infected millions of people worldwide. Thus, all technological gadgets that allow the fast detection of COVID-19 infection with high accuracy can offer help to healthcare professionals. This study is purposed to explore the effectiveness of artificial intelligence (AI) in the rapid and reliable detection of COVID-19 based on chest X-ray imaging. In this study, reliable pre-trained deep learning algorithms were applied to achieve the automatic detection of COVID-19-induced pneumonia from digital chest X-ray images.

Moreover, the study aims to evaluate the performance of advanced neural architectures proposed for the classification of medical images over recent years. The data set used in the experiments involves 274 COVID-19 cases, 380 viral pneumonia, and 380 healthy cases, which was collected from the available X-ray images on public medical repositories. The confusion matrix provided a basis for testing the post-classification model. Furthermore, an open-source library PyCM* was used to support the statistical parameters. The study revealed the superiority of Model VGG16 over other models applied to conduct this research where the model performed best in terms of overall scores and based-class scores. According to the research results, deep learning with X-ray imaging is useful in the collection of critical biological markers associated with COVID-19 infection. The technique is conducive for the physicians to make a diagnosis of COVID-19 infection. Meanwhile, the high accuracy of this computer-aided diagnostic tool can significantly improve the speed and accuracy of COVID-19 diagnosis.

## Introduction

The ongoing COVID-19 pandemic is a continuing coronavirus disease pandemic of 2019 (COVID-19) caused by extreme acute respiratory coronavirus syndrome 2 (SARS-CoV-2). Back in December 2019, the outbreak was first reported in Wuhan, China. Afterwards, it was declared a global public health emergency by the World Health Organization on January 30th, 2020, and then a pandemic on March 11th of the same year (WHO 2020, March 11).

Governments are scrambling to shut down borders, monitor communications closely, trace the infected, isolate the suspected cases. Nevertheless, the number of people getting affected by the virus is still soaring in most countries, and it is expected to continually increase before the medicine/vaccine is made available after numerous clinical trials. As for such a situation, the right situation must be understood to make the right decisions. Therefore, multiple testing is a priority to be addressed and has already started in most countries.

Nevertheless, it can be appreciated that these experiments are vitally important, but it takes time to be performed with absolute accuracy. It has the potential to pose a risk, which is because if the infected are not detected promptly, it leads to passing on the infection to others, which could lead to an explosive rise. It can result in devastation, especially in those densely-populated countries. The standard real-time COVID-19 test is called RT-PCR (Polymerase chain reaction) test that is purposed to determine the presence of antibodies against the virus (Corman et al. 2020). Furthermore, The molecular testing of respiratory samples is recommended for the identification and laboratory confirmation of COVID-19 infection. However, it takes much time and likely to produce false-negative outcomes, as well(Xie et al. 2020). Meanwhile, large-scale COVID-19 tests cannot be conducted in many developing countries due to its high cost. Where the immediate diagnosis depends on the symptoms appear.

Artificial Intelligence (AI) has recently been widely employed to accelerate biomedical research. AI was used in many applications, such as image detection, data classification, image segmentation, using deep learning approaches(Liu et al. 2019; Toğaçar et al. 2020). People infected with COVID-19 may suffer from pneumonia as the virus spreads to the lungs. Numerous profound learning studies have detected the disease using a chest X-ray imaging approach(Bougias et al. 2019; Jaiswal et al. 2019).

The remainder of this paper is organized as follows: Section 2 discusses the related works on COVID-19 classification. Section 3 describes our Methodology used in this study. The quantitive results and discussion is presented in Section 4. Section 5 presents the conclusion of this study.

### Related Work

Research COVID-19 classification literature has numerous methods with different datasets used in some study. Furthermore, many attempts are proposed some ready network with some changes in some cases to enhance the performance of the classification. Myriad works on COVID-19 classification are available, and we will briefly discuss the most relevant works. The American College of Radiology (ACR) advised against the use of CTs and X-rays as a first-line diagnostic or screen tool for COVID-19 diagnosis(the American College of Radiology 2020, March 22). Where they indicated that the images could only show signs of infection that may be due to other factors.

However, there have been plenty of studies where artificial intelligence was applied to test COVID-19 based on chest X-ray images(Afshar et al. 2020; Apostolopoulos & Mpesiana 2020; Farooq & Hafeez 2020; Hemdan et al. 2020; Khan et al. 2020; Li & Zhu 2020; Narin et al. 2020; Ozturk et al. 2020; Sethy & Behera 2020; Toraman et al. 2020; Ucar & Korkmaz 2020; Wang & Wong 2020).

Sethy and Behera(Sethy & Behera 2020) extracted the deep feature of nine convolutional neural network (CNN) pre-trained model and fed to SVM classifier individually. To choose the best classification model, statistical analysis was carried out. Their model achieved 95.38% of accuracy.

In(Narin et al. 2020) the authors experimented with three different CNN models (ResNet50, InceptionV3, and Inception-ResNetV2), and ResNet50 achieved the best accuracy of 98% for 2-class classification. Since they did not include pneumonia cases in their experiment, it is unknown how well their model would distinguish between COVID-19 and other pneumonia cases. In(Apostolopoulos & Mpesiana 2020) 224 approved COVID-19, 700 pneumonias, and 504 normal radiology images are used. They achieved a 98.75% performance for the 2-class and 93.48% performance for the 3-class problem.

The authores in (Hemdan et al. 2020) used deep learning models in X-ray images to diagnose COVID-19 and suggested a COVIDX-Net model consisting of seven CNN models. Wang and Wong(Wang & Wong 2020) presented a deep residual architecture called COVID-Net .it is one of the early works that has been done on COVID-19, which uses a deep neural network to classify chest X-ray images into three categories (COVID-19, Healthy, Non-COVID-19). COVID-Net achieved an accuracy of 92.4%. Ozturk et al.,(Ozturk et al. 2020) proposed a CNN model based on DarkNet architecture to detect and classify COVID-19 cases from X-ray images. Their model achieved binary and 3-class classification accuracy of 98.08% and 87.02%, respectively, of 125 COVID-19, 500 Pneumonia, and 500 healthy chest X-ray images. Farooq and Hafeez(Farooq & Hafeez 2020) presented COVID-ResNet for the classification of COVID-19 and three other infection types. COVID-ResNet was trained on a publicly available dataset.

In (Ucar & Korkmaz 2020) authors introduced a COVID-19 detection AI model, COVIDiagnosis-Net, based on deep SqueezeNet with Bayes optimization. The implemented deep learning model has obtained an accuracy performance of 98.3%. Asif Iqbal Khan(Khan et al. 2020) proposed an in-depth learning approach to detect COVID-19 cases from chest radiography images. The proposed method (CoroNet) is a convolutional neural network designed to identify COVID-19 cases using chest X-ray images. The experimental results indicated that the suggested model achieved an overall accuracy of 89.6%. Suat Toraman et al.,(Toraman et al. 2020) proposed a Convolutional CapsNet for the detection of COVID-19 disease by using chest X-ray images with capsule networks. Li and Zhu(Li & Zhu 2020) presented a novel mobile AI approach for CXR based COVID-19 screening called COVID-MobileXpert to be reliably deployed at mobile devices for point-of-care testing. Afshar et al., (Afshar et al. 2020) proposed a framework based on Capsule Network, known as the COVID-CAPS, for COVID-19 identification using X-ray images. The proposed COVID-CAPS achieved 95.7% accuracy, 90% sensitivity, 95.8% specificity, and 97% Area Under the Curve (AUC).

We noticed that some researchers combined the bacterial and viral pneumonia cases(Afshar et al. 2020). This combination is not appropriate and it will lead to inaccurate classification performance because the variance these combined two classes will be undistinguishable. If we tried to use only two classes e.g., normal and COVID-19 classes, some classification results cannot be distinguished between COVID-19 and viral pneumonia. The main problem is that, it cannot be distinguished between the viral pneumonia cases and COVID-19 cases. Therefore, we tried in this study to solve this problem by dividing the images in dataset into three classes they are normal, pneumonia cases and COVID-19.

In this study, we used COVID-19 chest images data set, viral pneumonia chest images, and healthy chest images, to evaluate the effectiveness of the state-of-the-art pre-trained Convolutional Neural Networks with regard to the automatic diagnosis of COVID-19 from chest X-rays. An automated detection of COVID-19 was proposed that applied pre-trained transfer models on Chest X-ray images based on a deep convolution neural network. A collection of 1034 chest X-rays images are stored and used for training and evaluation of the CNNs to achieve such a purpose. As the size of the COVID-19 related samples is small (274 images), transfer learning is considered to be a preferred strategy for training the deep CNNs. A combination of pre-trained models VGG16, DenseNet121, InceptionV3, InceptionResNetV2, MobileNet, DenseNet169, NASNetLarge, and Exception were employed to achieve a higher detection accuracy for a small X-ray dataset. Also, the current study will differ from previous studies by not relying on accuracy only during the model evaluation. Whereas, the accuracy is not considered an appropriate evaluation of the scarce and unequal data(Bi & Zhang 2018; Fernandes et al. 2019; McNee et al. 2006). Instead, the model was evaluated with several eligible measurements for such cases.

## Methodology

Challenge in COVID-19 recognition are vital, particularly when there are some viral pneumonia images. It’s difficult to differentiate between COVID-19 images and viral pneumonia images. Combining the viral pneumonia images with normal images also lead to misclassification results. Our method attempted to tackle this problem to identify COVID-19, viral pneumonia images and normal images by using pre-trained models with some modification in the top of these models. The pre-trained models directly learn its representation from the three classification of the dataset. Figure 1 shows our network, which is explained in detail as follows.

**Figure 1:**
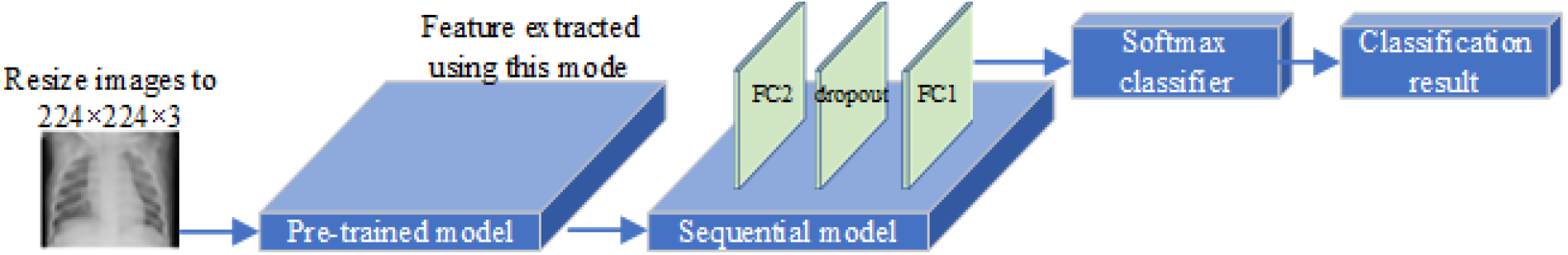
Outline of the methodology.

### Dataset and Input Preprocessing

As in previous studies, the dataset used in training is taken from multiple sources of X-rays(Cohen et al. 2020). the dataset made available online(Chowdhury et al. 2020) [22] for research purposes. The study was conducted by detailing the dataset for the confirmed COVID-19 cases, the cases of viral pneumonia infection (excluding COVID-19), and ‘healthy’ cases. Therefore, another dataset for healthy and viral pneumonia cases have been taken from Kaggle which also used in previous studes as in(Afshar et al. 2020; Apostolopoulos & Mpesiana 2020; Li & Zhu 2020; Narin et al. 2020; Sethy & Behera 2020; Wang & Wong 2020). The number of images in this dataset involves 274 COVID-19 cases, 380 viral pneumonia cases, and 380 healthy. The dataset was prepared and verified as reliable by reviewing it with chest specialists. Taking into account those cases of viral pneumonia should be free of any COVID-19 cases.

before being passed to pre-trained model for feature extraction, the images resized to the size of 224 × 224 × 3. CNN is capable of ignoring the insignificant variations in position. It searched for the patterns not only to a specific position of the image but also to moving patterns. During the training, the dataset divided into 70% for the training set and 30% for the validation set. Figure 2 shows examples of the dataset images used in training in this study.

**Figure 2:**
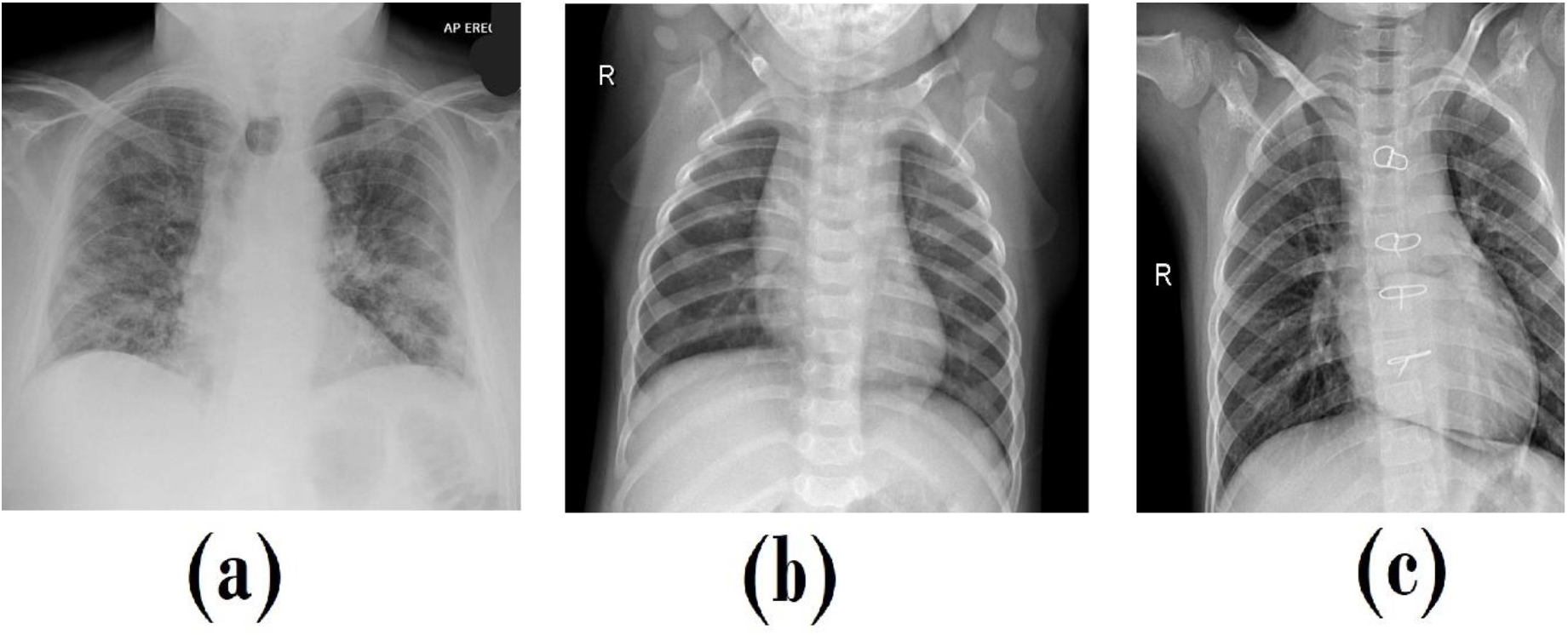
examples of the dataset images. (a) COVID-19, (b) Healthy, (c) Viral pneumonia.

### Pre-trained Model and Sequential Model

In this study, the transfer learning technique was applied that was introduced by using ImageNet data to resolve inadequate data and the time for preparation. The weights trained on ImageNet were downloaded for each model. The feature maps were treated as input size in the applied layers training process. Besides, since the convolution base was run on the small data set and the extracted features were taken as input, it worked in a highly efficient way. Thus, for fine-tuning, a brief description was made of the CNNs employed for automatic detection. Table 1 shows the CNNs applied for the classification function and criteria for transfer learning. The parameters of the fine-tuning were determined after several experiments. The number of possible choices was limitless; their contribution to improving efficiency could be explored in future research. The parameter called frozen layer refers to the number of untrainable layers starting from the top of the CNN, which is good because their weights are not expected to change during the process of model training. The last activation feature map in the pre-trained model provides us with the bottleneck features, which can then be flattened and fed into a fully connected deep neural network classifier. The other layers closer to the output features were trained to allow the extraction of more information from the late coevolutio nary layers. A dropout layer(Hinton et al. 2012) was added to prevent the occurrence of overlapping (Hawkins 2004) for neural networks using two hidden layers as depicted in Figure 1. The network is trained with a softmax classifier for 15 epochs by using an RMSprop optimizer(Tieleman & Hinton 2012) and learning rate of 0.00001 with the batch size set to 32. The result of training the pre-trained model produces a high feature representation of the images before these features passed to sequential model, while the sequential model used as a classifier with the last three layers including the softmax classifer. The consumed time to start the first epoch and the total training time for each model are shown in Table 1.

**Table 1:** The parameters of CNNs and computational time in seconds.

| Pre-trained models | Frozen layers | Time for training |  |
| --- | --- | --- | --- |
|  |  | first epoch | Total |
| InceptionV3 | 230 | 5 | 23 |
| Exception | 116 | 5 | 61 |
| InceptionResNetV2 | 682 | 14 | 84 |
| MobileNet | 67 | 2 | 30 |
| VGG16 | 18 | 3 | 45 |
| DenseNet169 | 575 | 12 | 68 |
| NASNetLarge | 819 | 20 | 146 |
| DenseNet121 | 407 | 9 | 65 |

## METRICS

In this paper, the performance of classification models to detect positive case of COVID-19 based on popular pre-trained models was evaluated. The proposed deep transfer learning models were trained separately using the Python Programming Language. All experiments were conducted with Tesla K80 GPU graphics card on Google Collaboratory with Windows 10 operating system. We used tenfold-cross-validation approach to assess the performance of our models.

The confusion matrix provides a common basis for the performance of classifiers to be evaluated. The literature on performance metrics based on confusion matrices is plentiful and diversified. in addition, it includes both frequent proposals for new statistics and the development of statistical models for their estimation. Here, we introduce an open-source Python library known as PyCM(Haghighi et al. 2018). It is not only a Python-based multi-class confusion matrix library purposed to support both the input and direct matrix data vectors but also a useful tool for evaluating the post-classification model that supports overall statistics parameters and class-based statistics parameters.

### Confusion matrix

A confusion matrix was introduced to analyze whether the prediction is consistent with the actual results. The confusion matrix is an effective method in assessing the classifier for its performance in classifying multi-class objects. This study focused on the general properties of the learning algorithm to address the problems with multi-class classification and measure quality with the confusion matrix. The instances in a predicted class represent each row of the matrix, while each column represents the instances in an actual class. The confusion matrix is regarded as one of the accurate measurements that provide more insight into the achieved validation accuracy. The three classes are investigated with the eight types of deep transfer learning. Nevertheless, such an assessment remains unclear, and the need for quantitative evaluation cannot be avoided(Carrillo et al. 2014).

### Overall performance statistics

The most used metric for reporting multi-class classification output is its sample accuracy (ACC), that is defined as the number of correct predictions in all classes k, as divided by the number of examples, n. Despite the simplicity of this concept, it is known that the assessment of performance using sample accuracy alone is likely to result in misinterpretation. This is because accuracy doesn’t account for the degree of class imbalance that can be present in a specific data set(Akbani et al. 2004; Brodersen et al. 2012; Chawla et al. 2002), which means that it can only be interpreted correctly in relation to a baseline accuracy based on the dataset. The average F1 scores per-class, known as F1_macro_, provide a commonly used way to solve the constraint mentioned above. Multi-class and multi-label classification problems are often assessed by the “F1_macro_” metric(Opitz & Burst 2019) and are computed as simple arithmetic means. As one of the dominant metrics in the remote sensing region, the Kappa coefficient(K_c_)(Cohen 1960) provides another solution to overcome the limitation on sample accuracy.

As with the F1_macro_ and K_c_, the class imbalance of the data is taken into account. Nonetheless, the number of errors may be invariant and do not necessarily represent one intuitively considered predictive power(Nishii & Tanaka 1999). An alternative is a macro-averaged accuracy (Acc_macro_)(Sammut & Webb 2011), which is defined as the arithmetic average of the partial accuracies of each class. Besides, other metrics were also computed, such as Overall Matthews Correlation Coefficient (MCC_overall_), which can be extended to multiple categories(Gorodkin 2004; Matthews 1975), Hamming loss (L_Hamming_), which is the fraction of wrong labels to the total number of labels, and the average true negative rate (TNR_macro_).

### Class-based performance statistics

In this section, the parameters for determining the COVID-19 class will be covered. Using one-versus all approach. we can determine the performance of classifiers with respect to only COVID-19 class. From a binary classification perspective some parameters must be computed. In this paper, accuracy(ACC), Matthews Correlation Coefficient (MCC), the area under curve(AUC), Geometric mean of specificity and sensitivity (GM), the harmonic mean of precision and sensitivity (F1), and true negative rate (TNR) of the model, are calculated. Among the retrieved situations, positive predictive value (PPV) also shoud be calculated which is the fraction of specific instances.

### Precision-recall metrics

Precision-Recall’s metric was also employed to estimate the quality of output for the classifier. Precision-Recall curves are deemed more informative when binary classifiers are evaluated on imbalanced data sets using such performance measures as precision and recall metrics. A high area under the curve of a precision-recall curve can be detected with either high precision or high recall, suggesting either a low false-positive rate or a low false-negative rate. The high scores for both indicate that the classifier is restoring not only accurate results (high precision) but also a majority of all positive results (high recall)—moreover, the higher f1-score, the more consistent of the classification model. Given the limitation on single metrics-precision, recall, and f1-score, an average precision score and precision-recall to each class were adopted to assess the overall capacity. Herein, average precision (AP) is involved in measuring the classifier for its accuracy using a weighted mean of precision achieved at each threshold. Furthermore, the output is binarized if the precision-recall curve and average precision were extended to multi-class classification. The precision-recall curve can be plotted along with iso-f1curves by considering each element of the label indicator matrix, which is regarded as a binary prediction (micro-averaging).

## Results And Discussion

The confusion matrix is shown in figure 3, based on the part of the dataset that used in validation during our training. Both false-negative and false-positive could affect medical decisions negatively. A false-positive result is produced when an individual is inaccurately assigned to a class, such as a healthy individual categorized as COVID-19 patient. False-negative results when an individual falling into a given class is excluded from such a group. As confirmed by the confusion matrix results, there was a consistency between the predicted and actual results, implying a better performance of the model in the classification of multi-class objects, as shown in Fig. 3.

**Figure 3:**
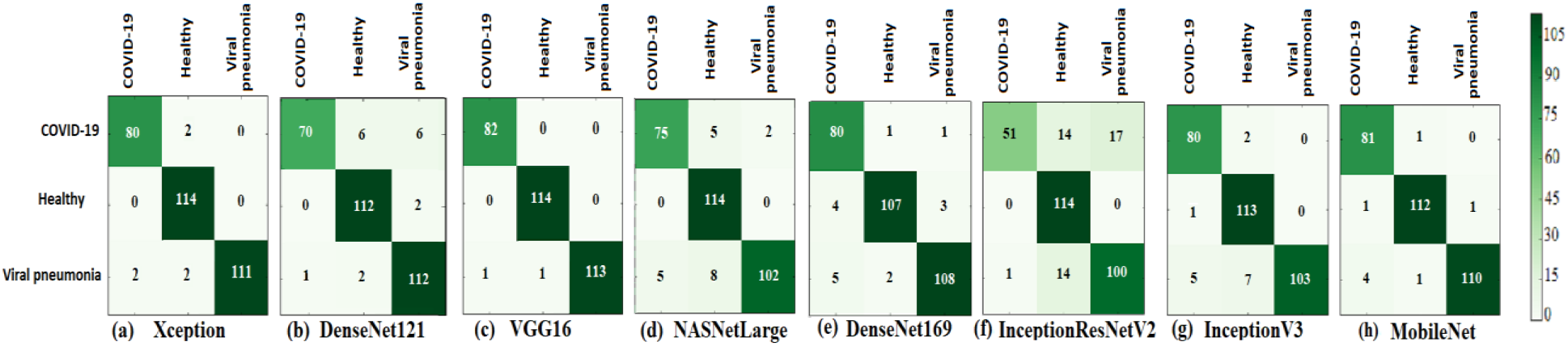
Confusion matrix of all deep learning models.

The model evaluated using the overall scores of the three classes. We also evaluated our model for class-based of COVID-19. Tables 2 and 3 show the overall and class-based parameters, respectively, as calculated from the confusion matrix while figure 4 and 5 demonstrate a comparison chart of overall and class-based parameters respectively.

**Table 2:** overall statistical parameters of different classification models (%)

| Classifiers | Acc<sub>macro</sub> | F1<sub>macro</sub> | L<sub>Hamming</sub> | K<sub>c</sub> | MCC<sub>overall</sub> | TNR<sub>macro</sub> |
| --- | --- | --- | --- | --- | --- | --- |
| Xception | **98.71** | **98.02** | **1.93** | **97.07** | **97.10** | **99.03** |
| DenseNet121 | 96.36 | 94.18 | 5.47 | 91.66 | 91.80 | 97.14 |
| VGG16 | **99.57\*** | **99.01 \*** | **0.64\*** | **99.03 \*** | **99.03\*** | **99.69\*** |
| NASNetLarge | 95.71 | 93.45 | 6.43 | 90.24 | 90.45 | 96.73 |
| DenseNet169 | 96.57 | 94.75 | 5.15 | 92.23 | 92.28 | 97.50 |
| InceptionResNetV2 | 90.14 | 83.80 | 14.79 | 77.23 | 78.31 | 92.22 |
| InceptionV3 | 96.79 | 95.17 | 4.82 | 92.70 | 92.86 | 97.60 |
| MobileNet | 98.29 | 97.34 | 2.57 | 96.10 | 96.13 | 98.76 |

**Table 3:** class-based Parameters of Different Classification Models (%)

| COVID-19 Classifiers | ACC | MCC | AUC | GM | F1 | TNR | PPV |
| --- | --- | --- | --- | --- | --- | --- | --- |
| Xception | 98.71 | 96.78 | 98.34 | 98.34 | 97.56 | 99.13 | 97.56 |
| DenseNet121 | 95.82 | 89.15 | 92.47 | 92.19 | 91.50 | 99.56 | 98.59 |
| VGG16 | **99.69 \*** | **99.17\*** | **99.78\*** | **99.78\*** | **99.39\*** | **99.56\*** | **98.80\*** |
| NASNetLarge | 96.14 | 89.98 | 94.64 | 94.59 | 92.59 | 97.82 | 93.75 |
| DenseNet169 | 96.46 | 91.27 | 96.82 | 96.81 | 93.57 | 96.07 | 89.89 |
| InceptionResNetV2 | 89.71 | 72.92 | 80.88 | 78.69 | 76.12 | 99.56 | 98.08 |
| InceptionV3 | 97.43 | 93.52 | 97.47 | 97.47 | 95.24 | 97.38 | 93.02 |
| MobileNet | 98.07 | 95.15 | 98.30 | 98.30 | 96.43 | 97.82 | 94.19 |

**Figure 4:**
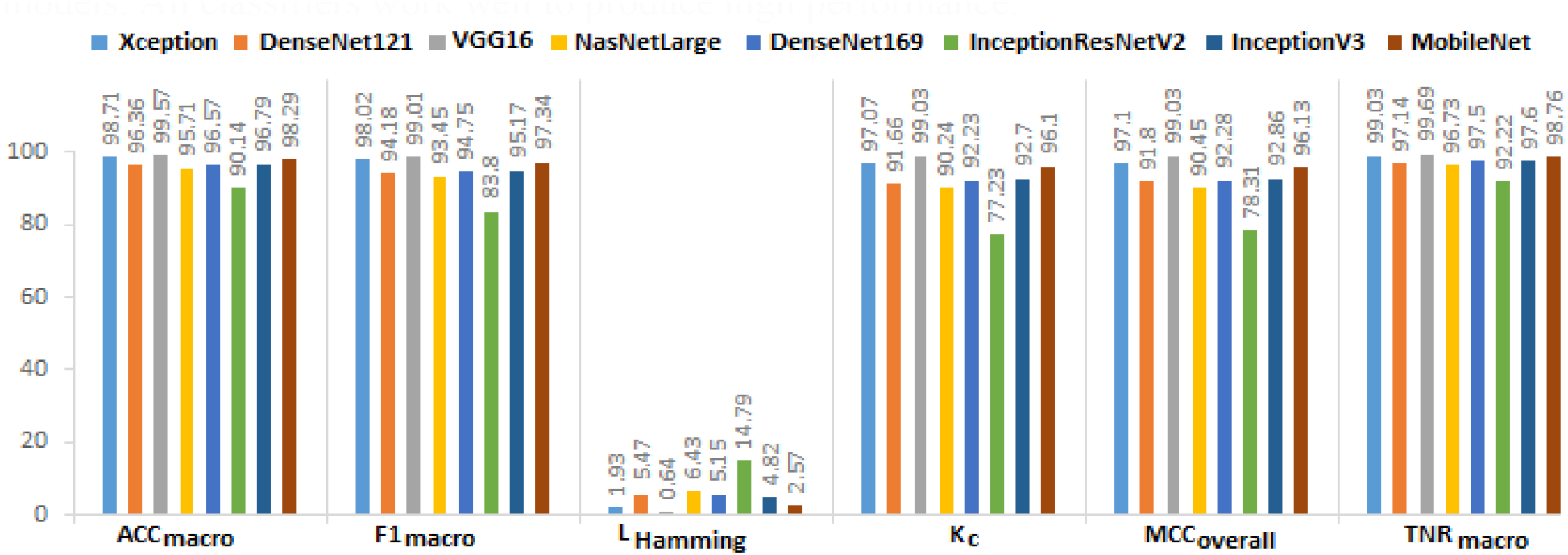
overall statistical parameters of different classification models.

**FIGURE 5:**
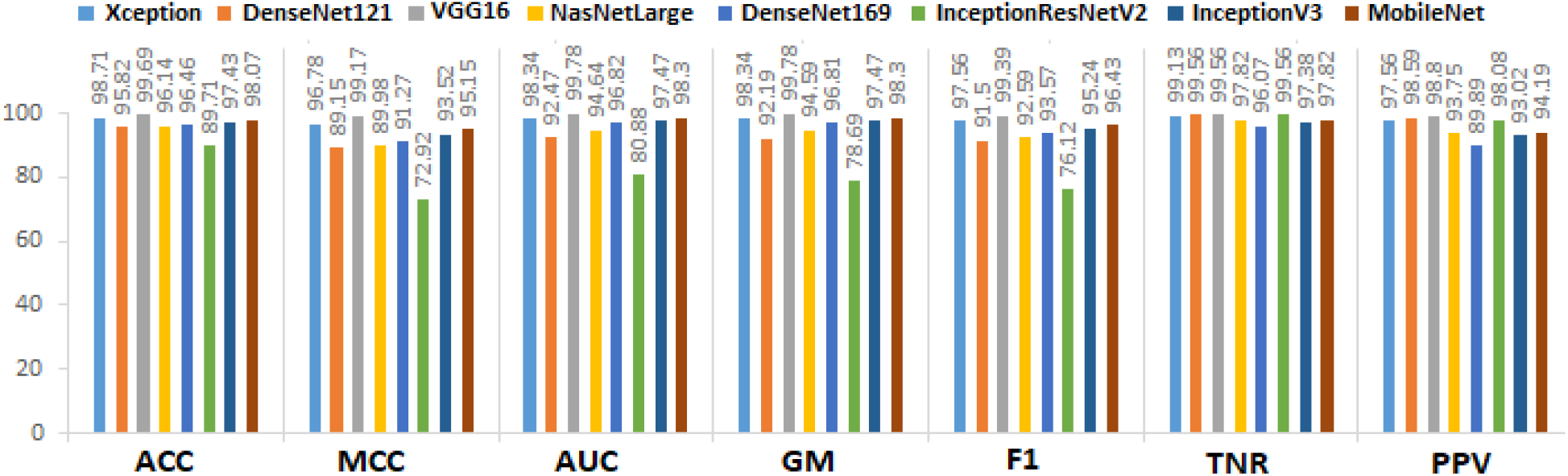
Class-based Parameters of Different Classification Models (%)

Table 2 shows the most satisfactory performance at a ACC_macro_ of 99.57%, F1_macro_ of 99.01%, L_Hamming_ of 0.64%, K_c_of 99.03%, MCC_overall_ of 99.03%, and TNR_macro_of 99.69% for the VGG16 classifier. The lowest performance values have been yielded at a ACC_macro_ of 90.14%, F1_macro_ of 83.80%, L_Hamming_ of 14.79%, K_c_ of 77.23%, MCC_overall_ of 78.31%, and TNR_macro_ of 92.22% for InceptionResNetV2. Consequently, the VGG16 model demonstrates its superiority to the other models. All classifiers work well to produce high performance.

Due to imbalanced data, the problem remains to determine which of these classifiers performs better in confirming COVID-19 infection. Any misdiagnosis may lead to severe consequences, especially concerning COVID-19 cases. As indicated by some of the results obtained from the confusion matrix, the classifier was capable of verifying all positive cases (COVID-19). However, it was wrong to consider some negative cases as positive cases. Besides, some of the classifiers were ineffective in verifying all positive cases, and they were indicated as negative cases. Therefore, it is necessary to calculate the parameters of the COVID-19 class, which is the target set for this paper. Table 3 shows the performance of the classifiers concerning only COVID-19 cases as our target class using one versus all approach, as supported in PyCM as well.

In addition to the parameters mentioned above, the detection model was also evaluated from the perspectives of precision-recall metrics. Figure 6 shows the average precision score for the classifiers, and Figure 7 shows the extension of the precision-recall curve to multi-classes. As revealed by the precision-recall curves shown in Fig. 6, the detection model proposed by us not only yielded a high average precision score in VGG16, Xception, and Mobilenet (AP = 0.99) (Fig. 6a,c,h) but also achieved a better performance in the detection of COVID-19 cases regarding the extension of the precision-recall curve to multi-classes (Fig. 7a,c,h).

**Figure 6:**
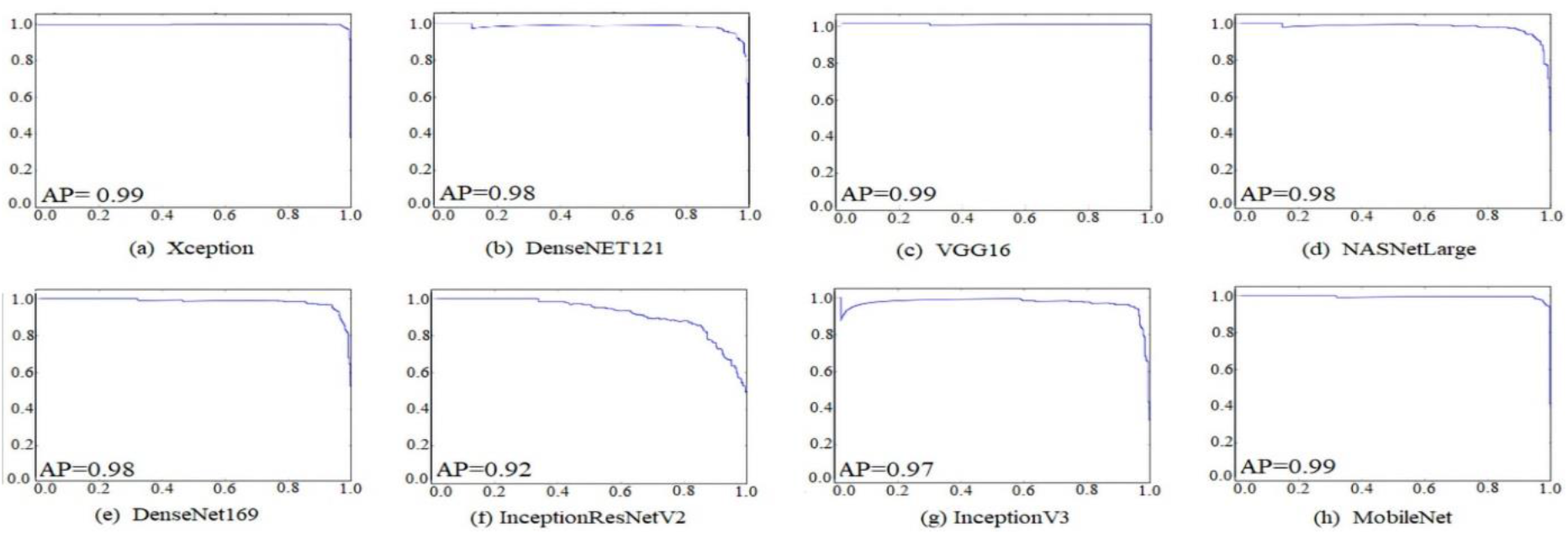
the average precision curves for the classifiers.

**Figure 7:**
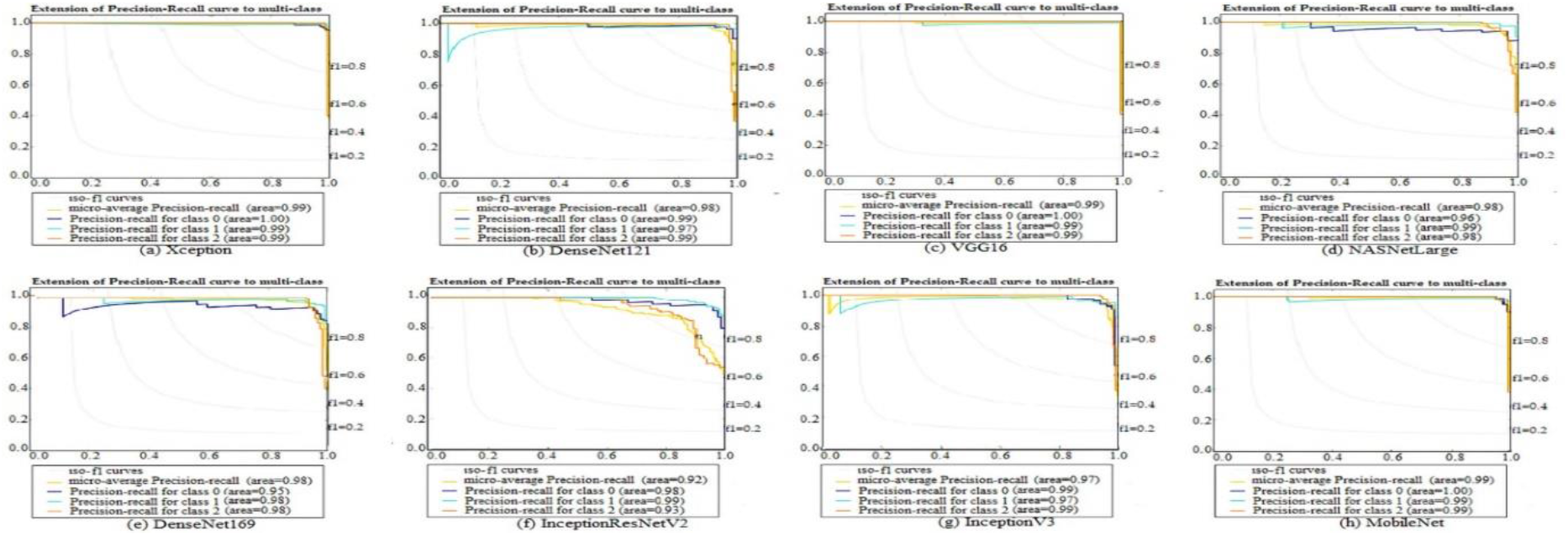
the precision-recall curve to multi-classes.

### Comparison between the State-of-the-Art methods

The AI techniques regarding the image classification approaches can help in early diagnose of the disease. Considering AI, CNN methods achieve better and faster results as compare to other classification methods. In this paper, a rapid, robust, and efficient COVID-19 diagnosis method is proposed. The proposed method performs the X-ray images into multi-class as healthy, viral pneumonia, and COVID-19. The general performance comparison of our study with the state-of-art methods is given in this section to evaluate the proposed CNN model. In the model evaluations, the related studies depend on the multi-class classification of the chest X-ray images with various AI techniques. Table 4 shows the comparison results with the related studies uses similar data sets. While Table 5 shows, the performance values of the listed studies are given in terms of COVID-19 class accuracy.

**Table 4:** The general comparison of the proposed method between state-of-the-art methods.

| Study | Method Used | Classes | ACC | TNR | F1 | MCC | K<sub>c</sub> |
| --- | --- | --- | --- | --- | --- | --- | --- |
| (Sethy & Behera 2020) | ResNet50+SVM | 3 |  |  | 95.52 | .9141 | .9076 |
| (Apostolopoulos & Mpesiana 2020) | VGG19 | 3 | 93.48 | 98.75 |  |  |  |
| (Apostolopoulos & Mpesiana 2020) | MobileNet v2 | 3 | 94.72 | 96.46 |  |  |  |
| (Wang & Wong 2020) | COVID-Net | 3 | 92.4 |  | 90 |  |  |
| (Ozturk et al. 2020) | DarkCOVIDNet | 3 | 87.02 | 92.18 | 87.37 |  |  |
| (Ucar & Korkmaz 2020) | Bayes-SqueezeNet | 3 | 98.3 | 99.1 | 98.3 | 97.4 |  |
| (Khan et al. 2020) | CORONET | 3 | 95 | 97.5 | 95.6 |  |  |
| (Toraman et al. 2020) |  | 3 | 84.22 | 91.79 | 84.21 |  |  |
| **This study** | **VGG16** | **3** | **99.57** | **99.68** | **99.36** | **99.03** | **99.03** |

**Table 5:** COVID-19 class comparison of the proposed method between the state-of-the-art methods.

| Study | Method Used | Classes | ACC | F1 | TNR |
| --- | --- | --- | --- | --- | --- |
| (Sethy & Behera 2020) | ResNet50+SVM | 2 | 95.38 |  | 93.47 |
| (Narin et al. 2020) | RESNET 50 | 2 | 98 | 98 | 100 |
| (Apostolopoulos & Mpesiana 2020) | VGG19 | 2 | 98.75 |  | 98.75 |
| (Hemdan et al. 2020) | VGG19 | 2 | 90 | 91 | 80 |
| (Ozturk et al. 2020) | DarkCOVIDNet | 2 | 98.08 | 96.51 | 95.3 |
| (Khan et al. 2020) | CoroNet | 2 | 99 | 98.5 | 98.6 |
| (Toraman et al. 2020) | CapsNet | 2 | 97.24 | 97.24 | 97.04 |
| **This study** | **VGG16** | **2** | **99.78** | **99.75** | **99.56** |

Sethy and Behera(Sethy & Behera 2020) employed the ResNet50 CNN model along with SVM for the detection of COVID-19 cases from chest X-ray images. CNN model acts as a feature extractor, and SVM serves the purpose of the classifier. Their model achieved an accuracy of 95.38% on the 2-class problem. Narin et al.,(Narin et al. 2020) used chest X-ray images coupled with the ResNet50 model to achieve a 98 % COVID-19 detection accuracy. Ioannis et al.,(Apostolopoulos & Mpesiana 2020) established the Deep Learning model using 224 confirmed COVID-19 images. Their model has achieved performance rates of 98.75%, and 93.48 % respectively for the two and three classes. Hemdan et al.,(Hemdan et al. 2020) introduced COVIDX-Net to detect COVID-19 in X-ray images. They got 90% accuracy by using 25 COVID-19 positive and 25 healthy images. Wang and Wong(Wang & Wong 2020) proposed a deep COVID-19 detection model (COVID-Net) that achieved 92.4% accuracy in the classification of four classes (healthy, non-COVID-19 pneumonia, and COVID-19). A CNN model proposed based on the DarkNet architecture by Ozturk et al.,(Ozturk et al. 2020) has achieved performance rates of 98.08% and 87.02%, respectively, for two and three classes. The COVIDiagnosis-Net model, which was proposed by Ferhat Ucar et al.,(Ucar & Korkmaz 2020) has achieved a performance accuracy of 98.3%. Asif Iqbal Khan(Khan et al. 2020) proposed a model called CORONET on chest X-ray images. CoroNet model managed to achieve an accuracy of 99% and 95% for 3-class and 2-class classification tasks respectively on a data set consisting of 224 COVID-19, 700 pneumonia, and 504 healthy X-ray images. Toraman S et al.,(Toraman et al. 2020) proposed a Convolutional CapsNetfor the detection of COVID-19 disease by using chest X-ray images with capsule networks. Their proposed method achieved an accuracy of 97.24% and 84.22%for binary class and multi-class, respectively. Amid the performance metrics that Tables 4 and 5 give, our model outperforms similar studies that use chest X-rays in the diagnosis of the COVID-19.

To the best of our knowledge, the proposed model reveals perfect and outstanding classification performance for the diagnosis COVID-19 with chest X-rays. A speedy and smooth implementation characterizes the work that we carried out. The promising and encouraging results of deep learning models in the detection of COVID-19 from radiography, images indicate that deep learning has a more significant role to play in fighting this pandemic soon.

## Conclusion

In this study, the overall and class-based parameters, respectively, were computed from the confusion matrix. According to the research results, the high performance can be achieved in multiclass-classification for all classifiers. Due to imbalanced data, however, the problem remains to identify which of these classifiers performs better in confirming COVID-19 cases. Any misdiagnosis may lead to severe consequences, especially concerning COVID-19 cases. Therefore, the parameters of the COVID-19 class were calculated in the study. The study revealed the superiority of Model VGG16 to other models applied in this research where the model achieved the highest values in terms of overall scores and based-class scores.

The study demonstrated that deep Learning with X-ray imaging might extract significant biological markers related to the COVID-19 disease. The technique is helpful to physicians in diagnosing COVID-19 patients. Meanwhile, the high accuracy of this computer-aided diagnostic tool can contribute to a significant improvement in the speed and accuracy of COVID-19 diagnosis.

For future studies, it is necessary to address other shortcomings. In particular, a more detailed analysis requires a more massive amount of patient data, especially those associated with COVID-19. Furthermore, such effective deep learning models as VGG16, and GoogLeNet, have been trained on more than a million images, which are barely available in the medical domain. Besides, there is a possibility that is training deep neural networks with limited data available results in over-fitting and hinders good generalization.

## Data Availability

All datasets used in the experiments were obtained from J. C. Monteral. (2020). COVID Chest X-ray Database (https://github.com/ieee8023/covid-chestxray-dataset), and COVID-19 Radiography Database (https://www.kaggle.com/tawsifurrahman/covid19-radiography-database)

https://doi.org/10.6084/m9.figshare.12863564.v1

